# Incidence, risk factors, and clinical symptom profile of reinfection during Omicron-dominated COVID-19 outbreak in Hong Kong: A retrospective cohort study

**DOI:** 10.1101/2024.03.12.24303945

**Authors:** Ziyang Huang, Jingyuan Luo, Haoran Li, Xingye Cheng, Jialing Zhang, Hiu To Tang, Hoi Ki Wong, Chun Hoi Cheung, Zhaoxiang Bian, Aiping Lyu, Liang Tian

**Affiliations:** Department of Physics, Hong Kong Baptist University, Hong Kong, China; Institute of Systems Medicine and Health Sciences, Hong Kong Baptist University, Hong Kong, China; School of Chinese Medicine, Hong Kong Baptist University, Hong Kong, China; Vincent V.C Woo Chinese Medicine Clinical Research Institute, School of Chinese Medicine, Hong Kong Baptist University, Hong Kong, China; Centre for Chinese Herbal Medicine Drug Development, Hong Kong Baptist University, Hong Kong SAR, China; Centre for Cancer & Inflammation Research, School of Chinese Medicine, Hong Kong Baptist University, Hong Kong, China; Institute of Computational and Theoretical Studies, Hong Kong Baptist University, Hong Kong, China; State Key Laboratory of Environmental and Biological Analysis, Hong Kong Baptist University, Hong Kong, China

**Keywords:** SARS-CoV-2, COVID-19, Omicron, reinfection, vaccination, risk factors

## Abstract

**Background:** Despite the World Health Organization’s declaration of the end of the COVID-19 pandemic, reinfection persists and continues to strain the global healthcare system. With the emergence of the most recent variant of SARS-CoV-2 named JN.1, retrospective analysis of epidemiological characteristics of previous cases involving the Omicron variant is essential to provide references for preventing reinfection caused by the ongoing new SARS-Cov-2 variants.

**Methods:** This retrospective cohort study included 6325 patients infected with SARS-CoV-2 during the Omicron-dominated outbreak (from December 2021 to May 2022) in Hong Kong. Statistical analysis was conducted to demonstrate the epidemiological characteristics and a logistic regression model was utilized to identify risk factors associated with reinfection.

**Results:** The Omicron reinfection incidence was 5.18% (n = 353). No significant difference was observed in receiving mRNA (BNT162b2) vaccine and inactivated (CoronaVac) vaccine between reinfection and non-reinfection groups (p>0.05). Risk factors were identified as female gender (p<0.001), longer infection duration (p<0.05), comorbidity of eyes, ear, nose, throat disease (p<0.01), and severe post-infection impact on daily life and work (p<0.05), while ≥70 years old (p<0.05) and vaccination after primary infection (p<0.01) were associated with a lower risk of reinfection. The prevalence of most symptoms after reinfection was lower than the first infection, except for fatigue.

**Conclusion:** No significant difference in mRNA (BNT162b2) vaccine and inactivated (CoronaVac) vaccine against reinfection. Post-infection vaccination could lower the risk of reinfection, which potentially inform the development of preventive measures including vaccination policies against potential new SARS-Cov-2 variants.

## 1. Introduction

Coronavirus disease 2019 (COVID-19) is a type of infectious disease caused by the coronavirus known as severe acute respiratory syndrome coronavirus 2 (SARS-CoV-2) [1,2]. As reported to WHO, there have been 710 million confirmed cases of COVID-19, including 6.9 million death cases as of September 26, 2023 [3–6]. There were different variants of SARS-CoV-2, including Alpha, Beta, Gamma, Delta, and Omicron, driving multiple infection waves worldwide [7]. The Omicron variant has rapidly become the dominant strain of the virus since its emergence due to immune escapability and high transmissibility [8].

The occurrence of reinfection was first confirmed in Hong Kong in August 2022 [9]. Although the World Health Organization (WHO) declared the end of the pandemic in May 2023 [10], the issue of reinfection persists. Previous studies have reported that Omicron reinfection has been occurring globally, with its incidence varying from different regions, such as 12.1% (Omicron BA.2 period) in Shanghai, China [11], 28.8% (Omicron BQ.1/BQ.1.1 period) in the U.S. [12], and 0.66% (Omicron BA.1/BA.2 period) in Kyoto, Japan [13]. As of January 5, 2024, the new variant named JN.1, an offspring of Omicron BA.2.86, was estimated to comprise approximately 62% of the currently circulating SARS-CoV-2 variants, continuously causing COVID-19 infections to increase globally [14]. It is crucial to retrospectively investigate previous cases of the Omicron variant to provide insights that can aid in developing preventive measures targeting potential new COVID-19 variants.

Apart from reinfection incidence, previous studies have identified several common risk factors associated with reinfection, such as gender, age, and vaccination status, with a particular focus on vaccination against reinfection [11,15–17]. However, other crucial factors that may influence reinfection risk, such as the infection duration of primary infection and post-infection impact on life, have not been thoroughly investigated in the current literature.

In Hong Kong, the Omicron variant was the dominant strain of the virus during the fifth wave of the outbreak, leading to over 2.8 million confirmed cases and 13,120 deaths from December 2021 to January 2023 [18]. The majority of cases were caused by Omicron BA.2 and its related sublineages (BA.2*) (85%) [19]. There were two types of COVID-19 vaccines available under the Government Vaccination Program in Hong Kong: the BNT162b2 mRNA Vaccine and the CoronaVac/Sinovac inactivated vaccine [20]. The symptom profiles in the acute stage and chronic stage of the Omicron outbreak in Hong Kong have been reported in our previous study [21,22]. Nevertheless, detailed studies on reinfection profiles were still limited. This study was based on a population of COVID-19 patients during the Omicron-dominated outbreak in Hong Kong, to systematically investigate the incidence, risk factors, as well as clinical symptom profile of COVID-19 reinfection.

## 2. Methods

### 2.1 Study design and population

This retrospective cohort study was conducted at the Vincent V.C. Woo Chinese Medicine Clinical Research Institute, Hong Kong Baptist University (HKBU). The data was collected from electronic medical records and revisit records via telephone from HKBU. As shown in Figure 1, during the Omicron-dominated outbreak in Hong Kong (December 31, 2021 – May 6, 2022), a total of 6814 patients with confirmed COVID-19 during the Omicron-dominated outbreak in Hong Kong were included. By excluding participants with hospitalization history after infection, 6325 non-hospitalized subjects with primary COVID-19 infection who received consultation services with complete electronic records from the university and revisit information were accounted for in the subsequent analysis. Follow-up telephone visits were conducted to assess the health status, including reinfection status by Chinese medicine physicians (CMPs) and research assistants (RAs) between November 21, 2022, and January 20, 2023. Primary infection and reinfection with SARS-CoV-2 were confirmed via polymerase chain reaction (PCR) or rapid antigen test (RAT). The data was collected under the Compulsory Testing Notice imposed by the Hong Kong Government from October 11, 2021 to December 29, 2022, which stated that individuals who have had close contact with confirmed cases of COVID-19 in 2019 but have not exhibited any symptoms of the disease are required to undergo mandatory quarantine at designated quarantine centers [23,24], minimizing the possibility of undetected reinfection among individuals who may be unaware of their reinfection status. The collected data consisted of reinfection status, demographic information, medical history, COVID-19 vaccination information, and post-infection symptoms.

**Figure 1.**
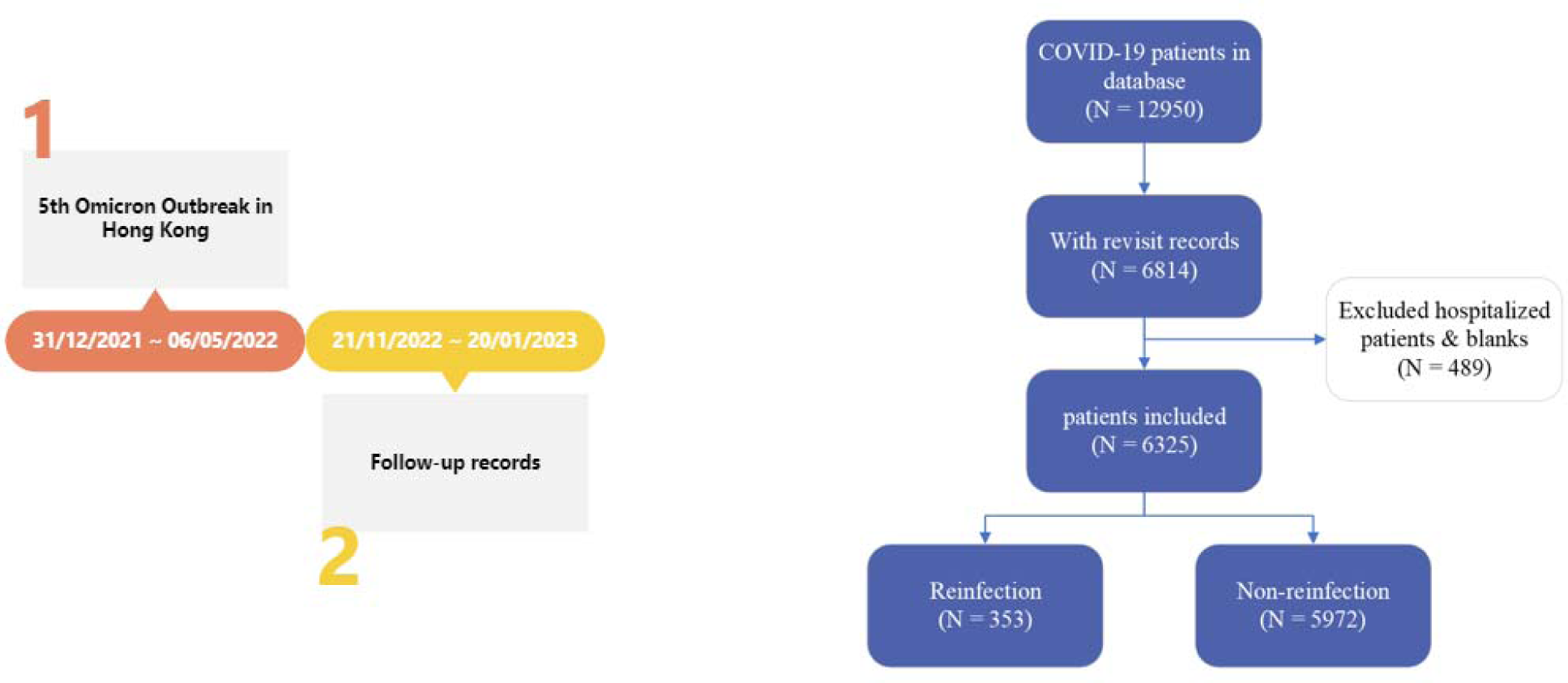
The flowchart of the data selection process.

### 2.2 Outcome

The incidence of COVID-19 reinfection was the primary outcome. The definition of reinfection from the CDC was applied in this study: an individual being infected by the SARS-Cov-2 virus, recovering, and then becoming infected again [25]. The investigated factors were as follows: gender, age, Body Mass Index (BMI), vaccination doses (incomplete vaccination: <3 doses; complete vaccination: ≥3 doses), vaccine types (BNT162b2, CoronaVac, and others), vaccination after primary infection, infection duration (measured in days, from the first positive to the first negative test results by PCR/RAT, comorbidities, treatment for acute stage of illness within four weeks after primary infection & corresponding satisfaction of acute treatment, new disease development after primary infection (new disease types were listed in Table S1), and self-reported post-infection impact on normal life and work in three levels: no impact, acceptable impact (mild to moderate), and severe impact. Fifteen clinical symptoms occurring after primary infection and reinfection were recorded.

### 2.2 Statistical analysis

For continuous variables, normally and non-normally distributed variables were expressed as mean (standard deviation, SD) and median (interquartile range, IQR) after testing normality, respectively, while the categorical variables were reported as frequencies and percentages (n%). Student’s t-test or Mann-Whitney U test was used for continuous variables as appropriate, while the Chi-squared test was utilized for categorical variables. To reduce bias arising from the unequal sample sizes between the reinfection group and non-reinfection group, we employed propensity score matching (PSM) [26], a statistical matching technique commonly utilized in observational studies to achieve a balanced distribution of characteristics between control and treatment groups, aiming for a 1:1 ratio of sample size, with propensity scores estimated using a logistic regression model (Figure 2). The associations between the risk factors and reinfection were assessed by logistic regression models, expressed as adjusted odd ratios (aOR) and unadjusted odd ratios (OR). Sex, age, BMI, vaccine dose, infection duration during primary infection, vaccination after primary infection, comorbidities, new disease development, and post-infection life impact were adjusted during the logistic regression analysis. All statistical tests were two-tailed, and the significance level was defined as *p* value < 0.05. Blank values were excluded from the analysis.

**Figure 2.**
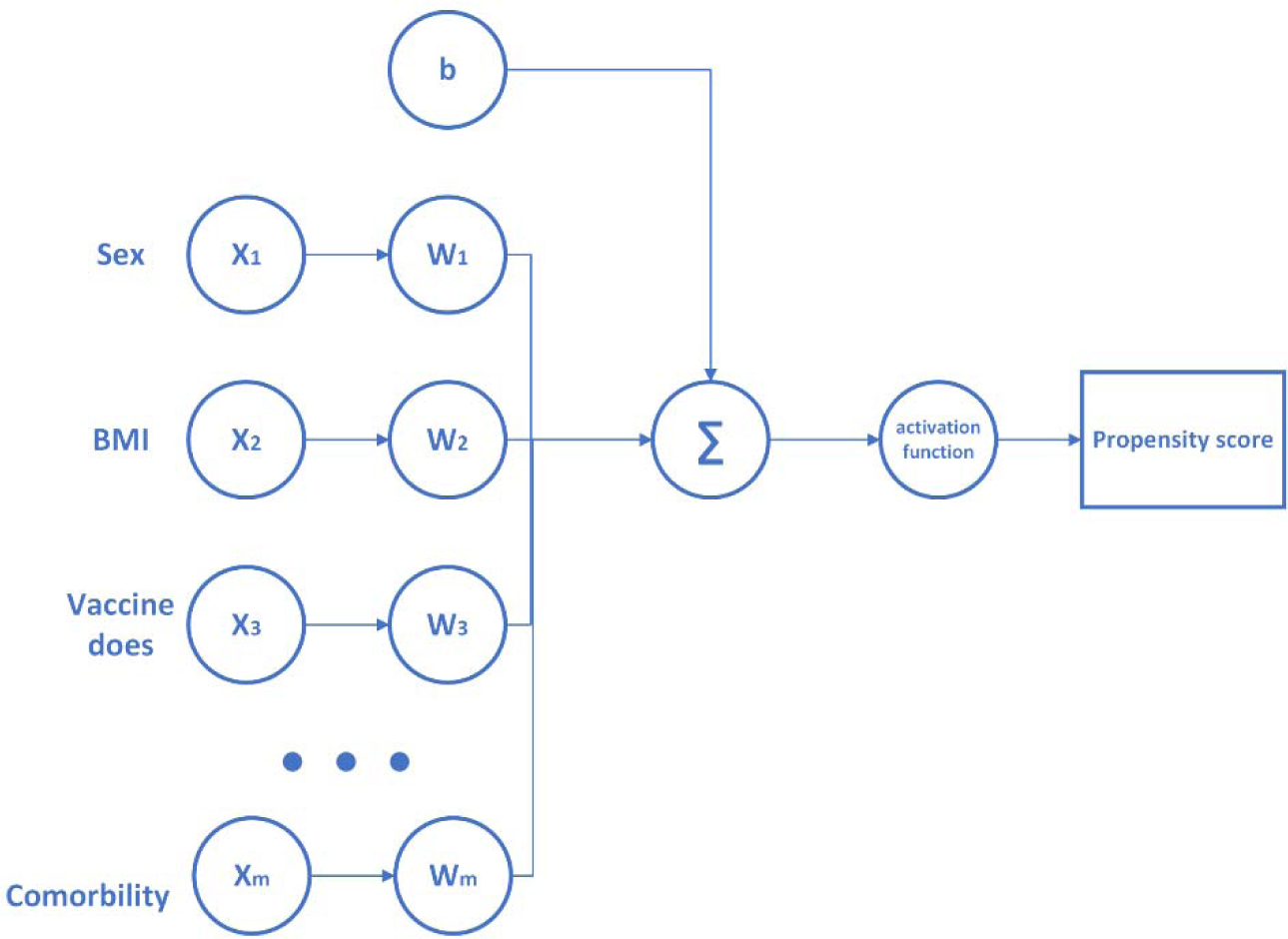
Demonstration of propensity score matching using logistic regression. Taking the age factor as an example, the rest of factors were used for matching.

## 3. Results

### 3.1 Cohort characteristics

The descriptive characteristics of the participant demography were shown in Table 1. The characteristic of the reinfection population were reported as follows: among reinfection patients, the median age was 44 (IQR: 35-58), with 76.77% female and 23.23% male. The median BMI was 22.8 (IQR: 20.7-25.9), and 17.28% were classified as obese. Furthermore, 44.76% of reinfection patients received at least three doses of vaccines, while 21.30% were vaccinated after primary infection. The median infection duration during the first infection was 7 days (IQR: 7-10). 29.75% of them had at least one comorbidity, and 3.68% developed new diseases within the investigated period. Regarding post-infection impact on daily life and work, 40.23%, 37.96%, and 9.92% of reinfection patients reported no impact, acceptable impact, and severe impact, respectively. For vaccine types (first vaccine dose), the proportions of mRNA (BNT162b2) and inactivated (CoronaVac) vaccines were 46.18% and 42.49%, respectively. Time intervals from first reinfection to primary infection (measured in days) were shown in Figure 3. The median time interval between primary infection and first reinfection was 279, with a minimum of 97 and a maximum of 333. The 95% confidence interval ranged from 155.97 to 311.72.

**Figure 3.**
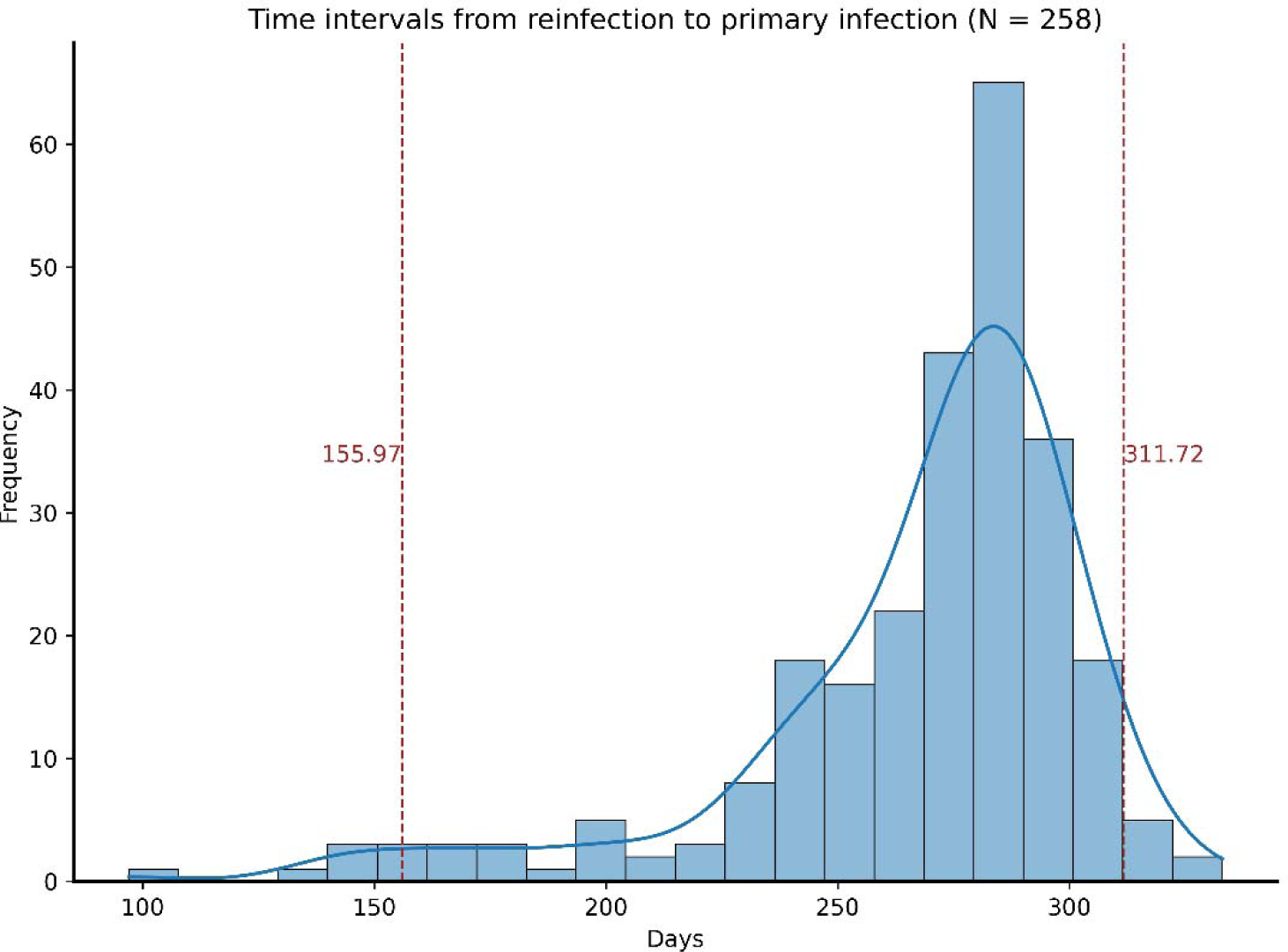
Distribution of time intervals from reinfection to primary infection. Only patients reinfected once (first reinfection) were included. Blank values were excluded.

**Table 1.**
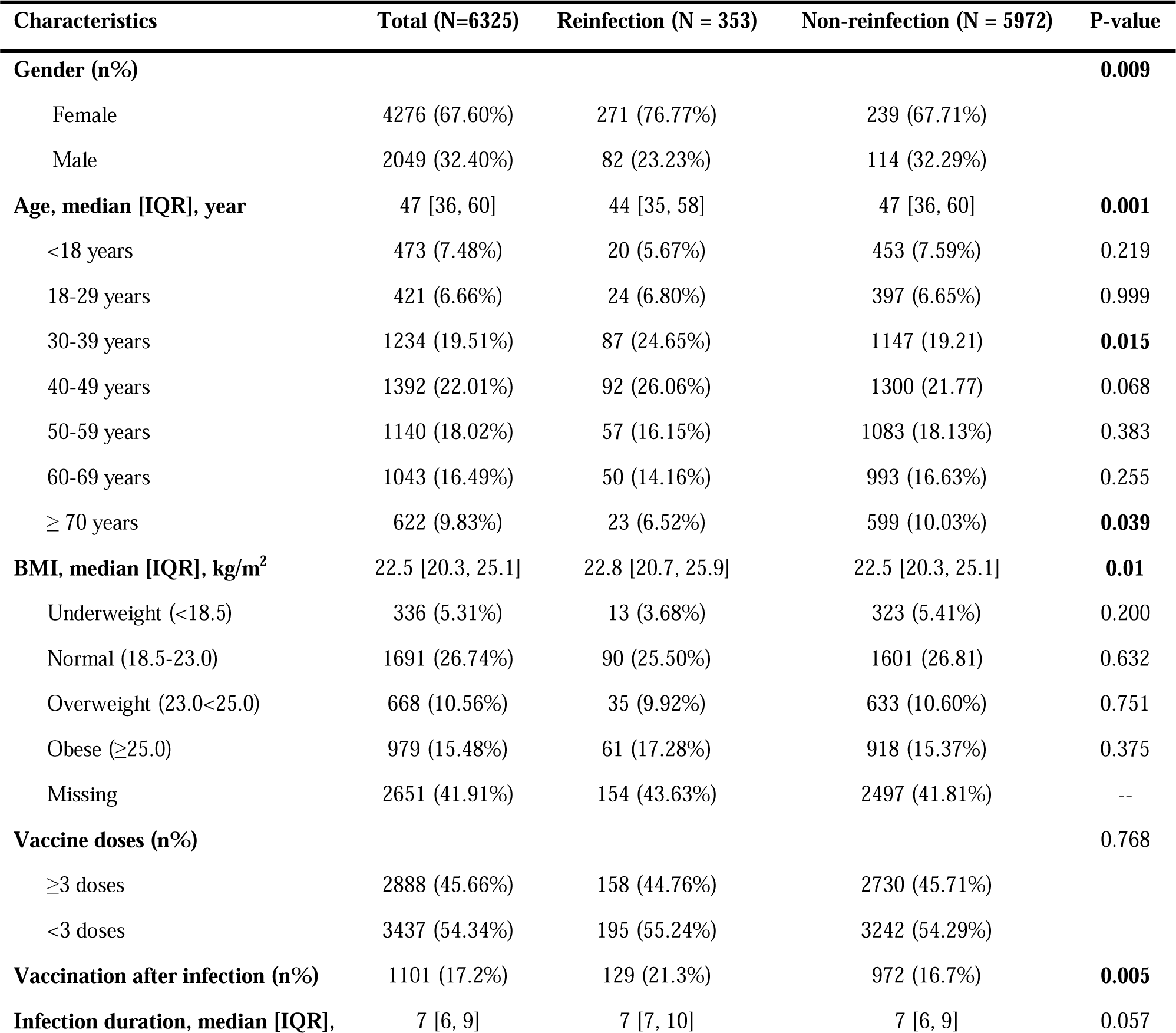
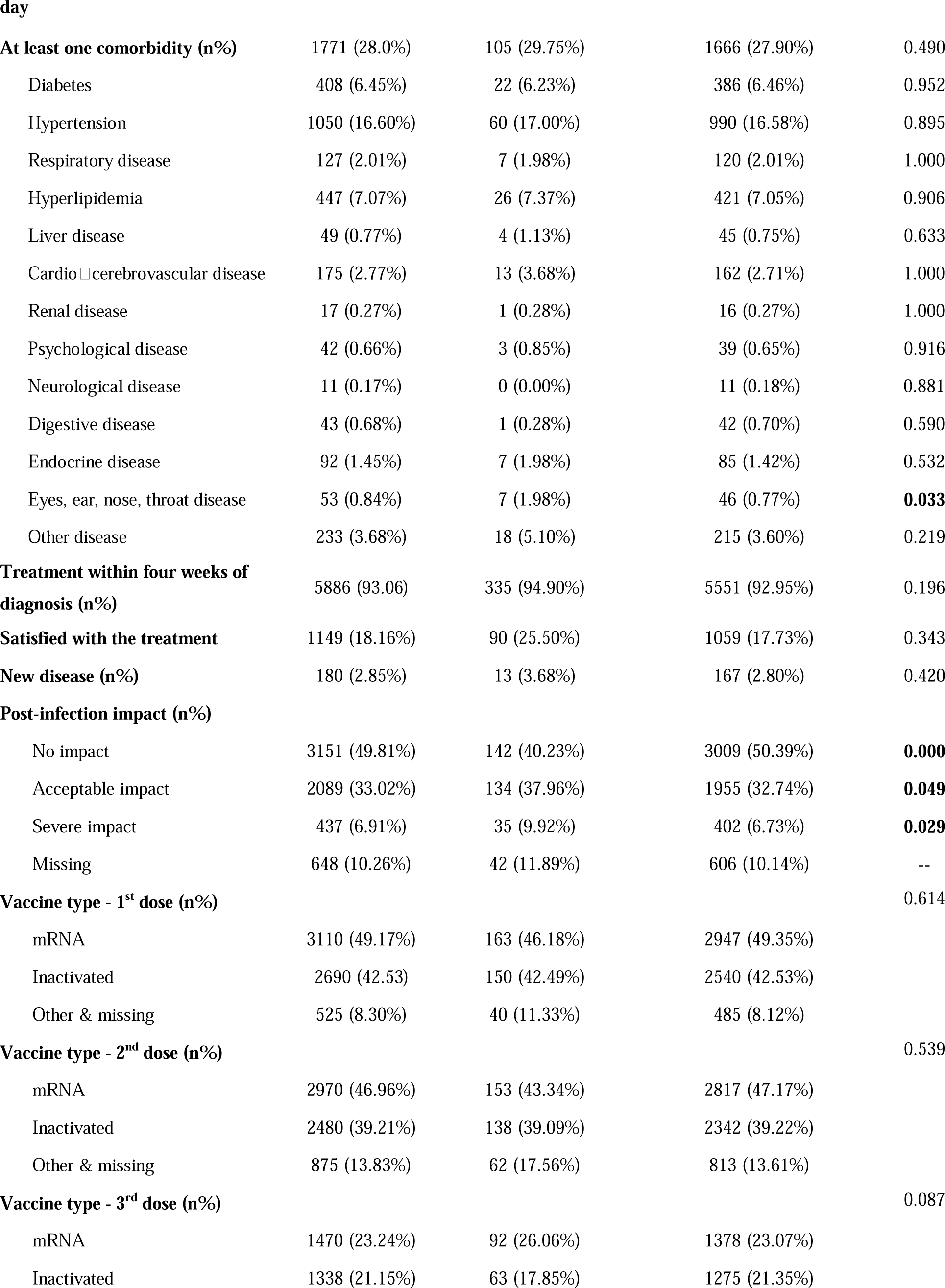
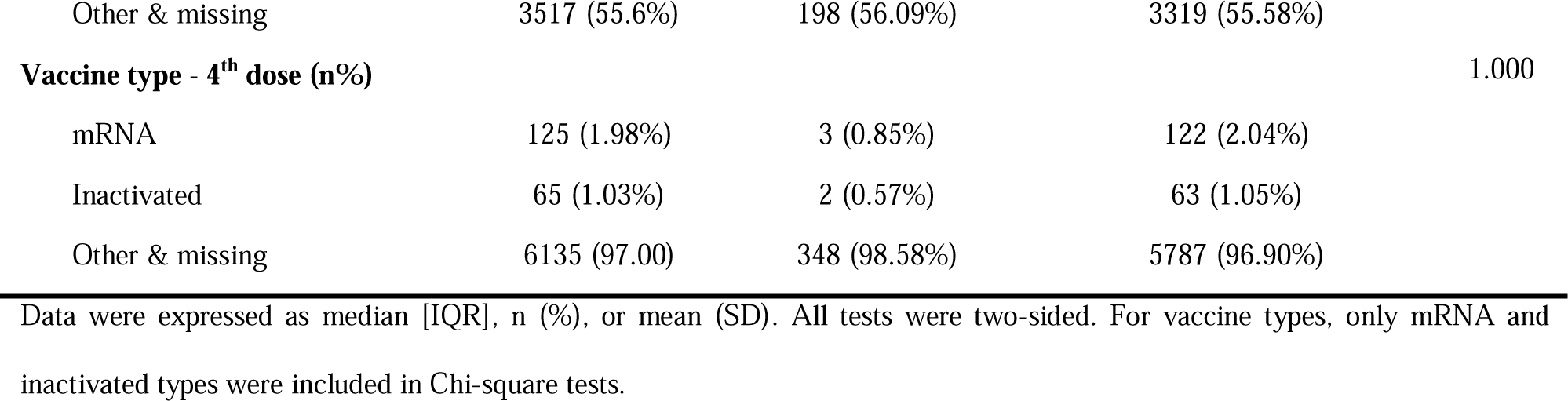
The overall demographic characteristics of the patients with follow-up records.

| Characteristics | Total (N=6325) | Reinfection (N = 353) | Non-reinfection (N = 5972) | P-value |
| --- | --- | --- | --- | --- |
| **Gender (n%)** |  |  |  | **0.009** |
| Female | 4276 (67.60%) | 271 (76.77%) | 239 (67.71%) |  |
| Male | 2049 (32.40%) | 82 (23.23%) | 114 (32.29%) |  |
| **Age, median [IQR], year** | 47 [36, 60] | 44 [35, 58] | 47 [36, 60] | **0.001** |
| <18 years | 473 (7.48%) | 20 (5.67%) | 453 (7.59%) | 0.219 |
| 18-29 years | 421 (6.66%) | 24 (6.80%) | 397 (6.65%) | 0.999 |
| 30-39 years | 1234 (19.51%) | 87 (24.65%) | 1147 (19.21) | **0.015** |
| 40-49 years | 1392 (22.01%) | 92 (26.06%) | 1300 (21.77) | 0.068 |
| 50-59 years | 1140 (18.02%) | 57 (16.15%) | 1083 (18.13%) | 0.383 |
| 60-69 years | 1043 (16.49%) | 50 (14.16%) | 993 (16.63%) | 0.255 |
| ≥ 70 years | 622 (9.83%) | 23 (6.52%) | 599 (10.03%) | **0.039** |
| **BMI, median [IQR], kg/m<sup>2</sup>** | 22.5 [20.3, 25.1] | 22.8 [20.7, 25.9] | 22.5 [20.3, 25.1] | **0.01** |
| Underweight (<18.5) | 336 (5.31%) | 13 (3.68%) | 323 (5.41%) | 0.200 |
| Normal (18.5-23.0) | 1691 (26.74%) | 90 (25.50%) | 1601 (26.81) | 0.632 |
| Overweight (23.0<25.0) | 668 (10.56%) | 35 (9.92%) | 633 (10.60%) | 0.751 |
| Obese (≥25.0) | 979 (15.48%) | 61 (17.28%) | 918 (15.37%) | 0.375 |
| Missing | 2651 (41.91%) | 154 (43.63%) | 2497 (41.81%) | -- |
| **Vaccine doses (n%)** |  |  |  | **0.768** |
| ≥3 doses | 2888 (45.66%) | 158 (44.76%) | 2730 (45.71%) |  |
| <3 doses | 3437 (54.34%) | 195 (55.24%) | 3242 (54.29%) |  |
| **Vaccination after infection (n%)** | 1101 (17.2%) | 129 (21.3%) | 972 (16.7%) | **0.005** |
| **Infection duration, median [IQR],** | 7 [6, 9] | 7 [7, 10] | 7 [6, 9] | 0.057 |
| **At least one comorbidity (n%)** | 1771 (28.0%) | 105 (29.75%) | 1666 (27.90%) | 0.490 |
| Diabetes | 408 (6.45%) | 22 (6.23%) | 386 (6.46%) | 0.952 |
| Hypertension | 1050 (16.60%) | 60 (17.00%) | 990 (16.58%) | 0.895 |
| Respiratory disease | 127 (2.01%) | 7 (1.98%) | 120 (2.01%) | 1.000 |
| Hyperlipidemia | 447 (7.07%) | 26 (7.37%) | 421 (7.05%) | 0.906 |
| Liver disease | 49 (0.77%) | 4 (1.13%) | 45 (0.75%) | 0.633 |
| Cardio□cerebrovascular disease | 175 (2.77%) | 13 (3.68%) | 162 (2.71%) | 1.000 |
| Renal disease | 17 (0.27%) | 1 (0.28%) | 16 (0.27%) | 1.000 |
| Psychological disease | 42 (0.66%) | 3 (0.85%) | 39 (0.65%) | 0.916 |
| Neurological disease | 11 (0.17%) | 0 (0.00%) | 11 (0.18%) | 0.881 |
| Digestive disease | 43 (0.68%) | 1 (0.28%) | 42 (0.70%) | 0.590 |
| Endocrine disease | 92 (1.45%) | 7 (1.98%) | 85 (1.42%) | 0.532 |
| Eyes, ear, nose, throat disease | 53 (0.84%) | 7 (1.98%) | 46 (0.77%) | **0.033** |
| Other disease | 233 (3.68%) | 18 (5.10%) | 215 (3.60%) | 0.219 |
| **Treatment within four weeks of diagnosis (n%)** | 5886 (93.06) | 335 (94.90%) | 5551 (92.95%) | 0.196 |
| **Satisfied with the treatment** | 1149 (18.16%) | 90 (25.50%) | 1059 (17.73%) | 0.343 |
| **New disease (n%)** | 180 (2.85%) | 13 (3.68%) | 167 (2.80%) | 0.420 |
| **Post-infection impact (n%)** |  |  |  |  |
| No impact | 3151 (49.81%) | 142 (40.23%) | 3009 (50.39%) | **0.000** |
| Acceptable impact | 2089 (33.02%) | 134 (37.96%) | 1955 (32.74%) | **0.049** |
| Severe impact | 437 (6.91%) | 35 (9.92%) | 402 (6.73%) | **0.029** |
| Missing | 648 (10.26%) | 42 (11.89%) | 606 (10.14%) | -- |
| **Vaccine type - 1<sup>st</sup> dose (n%)** |  |  |  | 0.614 |
| mRNA | 3110 (49.17%) | 163 (46.18%) | 2947 (49.35%) |  |
| Inactivated | 2690 (42.53) | 150 (42.49%) | 2540 (42.53%) |  |
| Other & missing | 525 (8.30%) | 40 (11.33%) | 485 (8.12%) |  |
| **Vaccine type - 2<sup>nd</sup> dose (n%)** |  |  |  | 0.539 |
| mRNA | 2970 (46.96%) | 153 (43.34%) | 2817 (47.17%) |  |
| Inactivated | 2480 (39.21%) | 138 (39.09%) | 2342 (39.22%) |  |
| Other & missing | 875 (13.83%) | 62 (17.56%) | 813 (13.61%) |  |
| **Vaccine type - 3<sup>rd</sup> dose (n%)** |  |  |  | 0.087 |
| mRNA | 1470 (23.24%) | 92 (26.06%) | 1378 (23.07%) |  |
| Inactivated | 1338 (21.15%) | 63 (17.85%) | 1275 (21.35%) |  |
| Other & missing | 3517 (55.6%) | 198 (56.09%) | 3319 (55.58%) |  |
| **Vaccine type - 4<sup>th</sup> dose (n%)** |  |  |  | 1.000 |
| mRNA | 125 (1.98%) | 3 (0.85%) | 122 (2.04%) |  |
| Inactivated | 65 (1.03%) | 2 (0.57%) | 63 (1.05%) |  |
| Other & missing | 6135 (97.00) | 348 (98.58%) | 5787 (96.90%) |  |

Data were expressed as median [IQR], n (%), or mean (SD). All tests were two-sided. For vaccine types, only mRNA and inactivated types were included in Chi-square tests.

### 3.2 The incidence of reinfection

The incidence of reinfection was 5.18% (n = 353). Among 353 reinfected patients, 65 (18.41%) of them have experienced reinfection twice (Figure S3). The propensity score matched comparison between the reinfection group and non-reinfection group was shown in Table 2. Compared with non-reinfection patients, the reinfection group has a higher percentage of females (271, 76.77% vs. 239, 67.71%, p<0.01), younger age population (44, 35-58 vs. 50, 38-62.25, p<0.005), higher BMI (22.8, 20.7-25.9 vs. 22.0, 20.0-24.6, p<0.05). Significant differences were found in post-infection impact on life and work between two groups (p<0.05), among which fewer reinfection patients reported no impact than non-reinfection patients (142, 44.65% vs. 176, 55.35%, p<0.05). Importantly, as shown in Table 3, there were no significant differences found in taking mRNA and inactivated vaccines between the two groups for either dose in four doses (p>0.05).

**Table 2.** Prevalence of reinfection using propensity score matched patient data.

| Characteristics | Reinfection (N = 353) | Non-reinfection (N = 353) | P-value |
| --- | --- | --- | --- |
| **Gender (n%)** |  |  | **0.009** |
| Female | 271 (76.77%) | 239 (67.71%) |  |
| Male | 82 (23.23%) | 114 (32.29%) |  |
| **Age, median [IQR], year** | 44 [35, 58] | 50 [38, 62.25] | **0.001** |
| **BMI, median [IQR], kg/m<sup>2</sup>** | 22.8 [20.7, 25.9] | 22.0 [20.0, 24.6] | **0.01** |
| **Vaccine dose, median [IQR], dose** | 2 [2, 3] | 2 [2, 3] | $>0.05$ |
| **At least one comorbidity (n%)** | 105 (29.75%) | 91 (25.78%) | $>0.05$ |
| **Infection duration, median [IQR], day** | 7 [7, 10] | 7 [6.75, 10] | $>0.05$ |
| **Post-infection impact (n%)** |  |  | **0.036** |
| No impact | 142 (44.65%) | 176 (55.35%) | **0.013** |
| Acceptable impact | 134 (43.09%) | 118 (37.11%) | $>0.05$ |
| Severe impact | 35 (11.25%) | 24 (7.55%) | $>0.05$ |
| **New disease development** | 13 (3.69%) | 13 (3.69%) | $>0.05$ |

The propensity score matching was performed among eight factors (bolded). During the matching process of each factor, the other seven factors were matched.

**Table 3.**
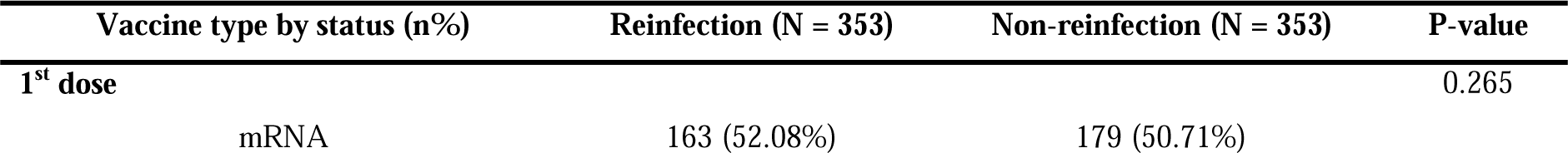
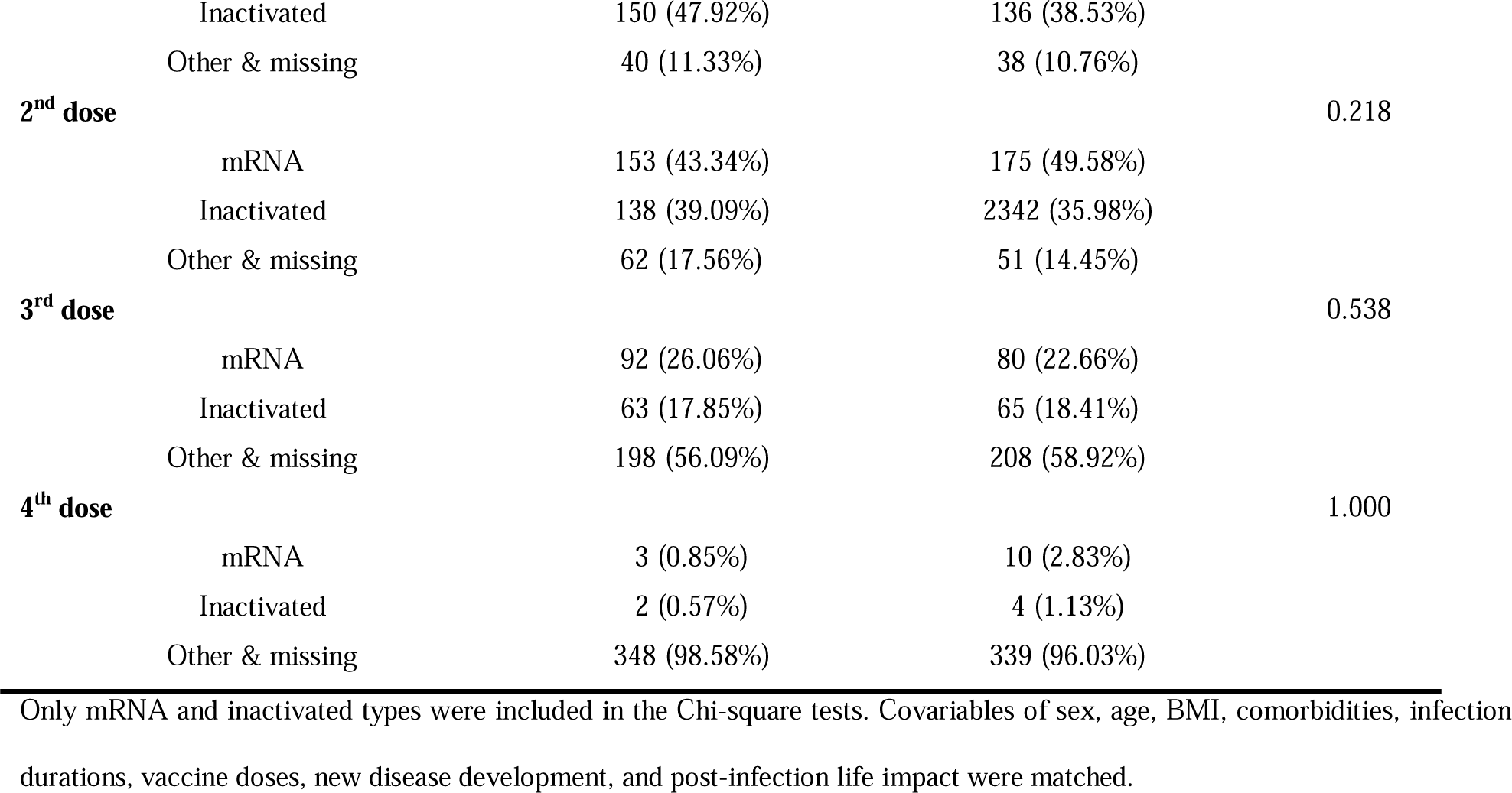
The propensity score matched comparison of vaccine types (mRNA vs. inactivated) of four vaccination status.

### 3.3 Risk factors associated with the incidence of reinfection

Table 4 presented the association between risk factors and reinfection using logistic regression. Several factors were identified to be associated with reinfection. Males exhibited a lower risk of reinfection than females (aOR: 0.619, 0.477-0.804, p<0.001), and individuals that were ≥70 years old had a lower reinfection risk than other age groups (aOR: 0.444, 0.231-0.852, p<0.05). A longer infection duration for primary infection was associated with a higher reinfection risk than a shorter infection duration (aOR: 1.030, 1.001-1.061, p<0.05). Vaccination after primary infection was significantly associated with a lower risk of reinfection (aOR: 0.708, 0.548-0.916, p<0.01). Having comorbidity of eyes, ear, nose, throat disease could increase the risk of reinfection (aOR: 2.983, 1.316-6.761, p<0.01). In terms of post-infection life impact, individuals who experienced either an acceptable impact (aOR: 1.316, 1.027-1.687, p<0.05) or severe impact (aOR: 1.651, 1.108-2.459, p<0.05), compared to those who reported no impact.

**Table 4.** Risk factors associated with the prevalence of reinfection.

| Factors | Unadjusted Odds Ratio | 95% CI | P-value | Adjusted Odds Ratio | 95% CI | P-value |
| --- | --- | --- | --- | --- | --- | --- |
| **Sex, (female as Ref.)** | 0.616 | [0.479, 0.793] | 0.000 | 0.619 | [0.477, 0.804] | **0.000** |
| **Age** |  |  |  |  |  |  |
| <18 years | 0.730 | [0.397, 1.342] | 0.311 | 0.877 | [0.471, 1.635] | 0.680 |
| 18-29 years | Ref. |  |  | Ref. |  |  |
| 30-39 years | 1.255 | [0.787, 2.000] | 0.340 | 1.163 | [0.726, 1.862] | 0.531 |
| 40-49 years | 1.171 | [0.737, 1.860] | 0.505 | 0.993 | [0.618, 1.595] | 0.977 |
| 50-59 years | 0.871 | [0.533, 1.422] | 0.580 | 0.689 | [0.414, 1.147] | 0.152 |
| 60-69 years | 0.833 | [0.505, 1.374] | 0.474 | 0.646 | [0.378, 1.104] | 0.110 |
| ≥ 70 years | 0.635 | [0.354, 1.141] | 0.129 | 0.444 | [0.231, 0.852] | **0.015** |
| **BMI** |  |  |  |  |  |  |
| Underweight (<18.5) | 0.716 | [0.395, 1.296] | 0.270 | 0.715 | [0.391, 1.307] | 0.276 |
| Normal (18.5 < 23.0) | Ref. |  |  | Ref. |  |  |
| Overweight (23.0 < 25.0) | 0.984 | [0.659, 1.469] | 0.936 | 1.097 | [0.730, 1.648] | 0.657 |
| Obese (≥25.0) | 1.182 | [0.846, 1.652] | 0.328 | 1.290 | [0.912, 1.825] | 0.150 |
| Missing BMI | 1.097 | [0.840, 1.434] | 0.497 | 1.233 | [0.936, 1.624] | 0.137 |
| **Vaccine dose** |  |  |  |  |  |  |
| <3 doses | Ref. |  |  | Ref. |  |  |
| ≥3 doses | 0.788 | [0.447, 1.387] | 0.409 | 1.071 | [0.853, 1.346] | 0.554 |
| **Infection duration** | 1.031 | [1.003, 1.060] | 0.029 | 1.030 | [1.001, 1.061] | **0.046** |
| Missing | 1.872 | [0.239, 14.674] | 0.551 | 1.803 | [0.225, 14.421] | 0.579 |
| **Vaccination after infection** |  |  |  |  |  |  |
| NO | Ref. |  |  | Ref. |  |  |
| YES | 0.697 | [0.545, 0.892] | 0.003 | 0.708 | [0.548, 0.916] | **0.009** |
| Missing | 0.531 | [0.313, 0.900] | 0.019 | 0.808 | [0.537, 1.217] | 0.308 |
| **At least one comorbidity, n (%)** |  |  |  |  |  |  |
| Diabetes | 0.919 | [0.560, 1.508] | 0.738 | 1.067 | [0.637, 1.787] | 0.805 |
| Hypertension | 1.022 | [0.742, 1.408] | 0.894 | 1.322 | [0.924, 1.890] | 0.126 |
| Respiratory disease | 0.955 | [0.441, 2.067] | 0.908 | 0.873 | [0.400, 1.907] | 0.734 |
| Hyperlipidemia | 1.048 | [0.657, 1.671] | 0.845 | 1.145 | [0.701, 1.868] | 0.589 |
| Liver disease | 1.544 | [0.551, 4.327] | 0.409 | 1.767 | [0.624, 5.003] | 0.284 |
| Cardio-cerebrovascular disease | 1.321 | [0.766, 2.280] | 0.317 | 1.636 | [0.929, 2.881] | 0.088 |
| Renal disease | 0.993 | [0.130, 7.600] | 0.995 | 1.396 | [0.179, 10.916] | 0.750 |
| Psychological disease | 1.177 | [0.359, 3.865] | 0.788 | 1.137 | [0.342, 3.778] | 0.834 |
| Endocrine disease | 1.392 | [0.637, 3.038] | 0.407 | 1.236 | [0.560, 2.729] | 0.600 |
| Eyes, ear, nose, throat disease | 2.643 | [1.182, 5.910] | 0.018 | 2.983 | [1.316, 6.761] | **0.009** |
| **New disease development** | 1.329 | [0.748, 2.361] | 0.333 | 1.073 | [0.594, 1.938] | 0.816 |
| **Post-infection impact** |  |  |  |  |  |  |
| No impact | Ref. |  |  | Ref. |  |  |
| Acceptable impact | 1.452 | [1.139, 1.852] | 0.003 | 1.316 | [1.027, 1.687] | **0.030** |
| Severe impact | 1.845 | [1.256, 2.709] | 0.002 | 1.651 | [1.108, 2.459] | **0.014** |
| Missing | 1.469 | [0.040, 0.056] | 0.034 | 1.522 | [1.057, 2.191] | 0.024 |

### 3.4 Symptom profiles of reinfection

Figure 4 shows the symptom profile of the first reinfection and primary infection of the same population (n = 278), which included the prevalence of 15 symptoms among those who were reinfected once. The top 5 most prevalent symptoms after reinfection were identified as follows: fatigue (34.17%), cough (33.09%), sputum (12.95%), headache (10.79%), and muscle pain (9.71%). Significant differences were observed in most of the symptoms between the reinfection group and the primary infection group. Among them, cough, sputum, dry throat, sore throat, itchy throat, abdominal distension, and diarrhea showed highly significant differences (p<0.001), while chest tightness, abdominal pain, and nausea exhibited significant differences (p<0.01). For those who had a primary infection and reinfection afterward, their symptom types experienced during reinfection were generally similar to those observed during the primary infection. The prevalence of most of the investigated symptoms after reinfection was lower than that observed during the primary infection.

**Figure 4.**
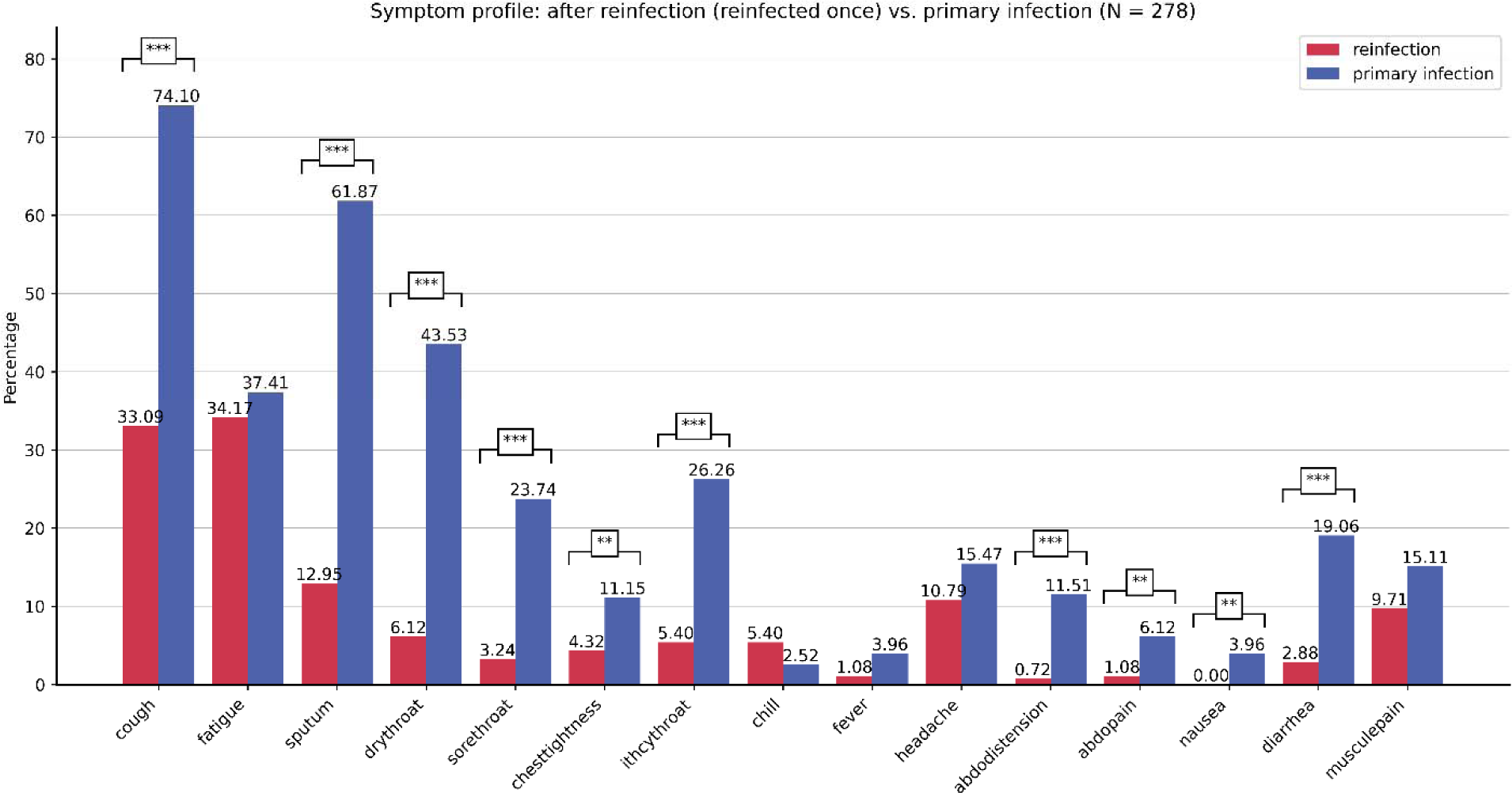
Symptom profile (the prevalence of 15 symptoms) of the first reinfection vs. primary infection. Patients with one reinfection was included in this plot, while patients had two reinfections were excluded. “***” means p<0.001; “**” means p<0.01.

## 4. Discussion

To our knowledge, this study investigated the incidence, risk factors, and clinical symptom profile of

Omicron reinfection in Hong Kong for the first time, with a large sample size and an overall age distribution. Based on our results, the reinfection incidence was 5.18% during the Omicron outbreak in Hong Kong, which was different from other studies. A reinfection rate of 11.5% was reported in Iceland during the Omicron period [27]. A study conducted in Korea reported a reinfection rate of 3% during the Omicron BA.2 period [28]. The reinfection incidence was 0.66% (Omicron BA.1-dominated period) in Kyoto, Japan [13]. In general, the reinfection rates from different areas reflect that the Omicron variant has been a persistent threat to the global healthcare system.

Our finding suggested that females were more likely to have reinfection, which aligned with the cases observed in primary infection in previous studies [22,29,30]. With respect to age, a plausible explanation for the increased susceptibility to reinfection among individuals over 70 years old could be their tendency to have fewer social interactions or lower exposure to high-risk environments compared to younger individuals, which may reduce their chances of encountering situations that lead to reinfection [13]. According to a report from the CDC, patients with a weakened immune system are more likely to be infected with COVID-19 for a more extended period [31]. Therefore, a longer duration of primary infection could increase the risk of reinfection, which may imply that individuals with a lower level of immune response could be more susceptible to reinfection. Additionally, comorbidity of ear, nose, throat disease may compromise immune barrier function in the nasal and pharyngeal regions, potentially increasing susceptibility to reinfections. Individuals who experienced a severe impact on life and work were at a higher risk of reinfection, which may also be attributed to residual immune deficiencies following primary infection recovery [31]. These hypotheses require further research to be confirmed.

Notably, vaccination after primary infection was associated with a lower risk of reinfection, which was in line with several other studies [32,33]. Our results also suggested no significant difference found in reinfection rates between the CoronaVac vaccine and the BNT162b2 vaccine. However, a previous study has reported that receiving mRNA vaccines leads to a lower reinfection rate than inactivated vaccines [34], while another study suggested that Combined vaccination may offer superior reinfection protection than either alone [35]. Considering the potential long-term persistence of the SARS-CoV-2 virus, individuals with relatively lower immune responses may benefit from a booster vaccination after primary infection, and further investigations were required to confirm the effects of different vaccine types.

Regarding the clinical symptom profile, the incidence of most reinfection symptoms was significantly lower than primary infection. This was consistent with a study reporting that 80.7% of the reinfection patients had mitigated or similar clinical symptoms compared with their first infection [11]. However, the prevalence of fatigue did not exhibit a significant difference. Fatigue, as one of the most common symptoms of long COVID, is prone to persist with repeated infections [21,36]. The lack of medications specifically targeting post-infection fatigue in COVID-19 patients requires further research.

### 4.1 Strengths and limitations

The strengths of our study are as follows. PSM was utilized in statistical analysis to balance the unequal sample sizes of the reinfection and non-reinfection groups, thereby making our results more reliable. In addition to demographic factors, several additional factors such as infection duration during primary infection, comorbidity of eyes, ear, nose, throat disease, and impact on life and work after primary infection were accounted for the first time in this study. Clinical symptoms of reinfection versus the first infection were analyzed in this study, providing a more comprehensive understanding of the reinfection profile.

There are several limitations in our study. First, the reinfection incidence might be underestimated as some reinfected individuals might not have been diagnosed. Second, follow-up records via telephone may introduce subjective bias and affect data reliability. Furthermore, the investigated population was regional and limited to Hong Kong, which may not represent the general population well.

## 5. Conclusion

In conclusion, this retrospective cohort study investigated the prevalence, risk factors, and symptom profile of COVID-19 reinfection during the Omicron-dominated outbreak in Hong Kong. No significant difference was found in the BNT162b2 vaccine and the CoronaVac vaccine. Populations with female gender, longer infection duration, having comorbidity of eyes, ear, nose, throat disease, having post-infection life impact, and no vaccination after primary infection were identified to be more susceptible to experience reinfection, while being ≥70 years old exhibited a lower reinfection risk. Fewer patients reported experiencing the majority of investigated clinical symptoms except fatigue during reinfection compared to their first infection. Notably, the finding of vaccination after infection can potentially inform the development of epidemic preventive measures, particularly policies regarding booster vaccination after infection, and ongoing updates for vaccines are also imperative to effectively combat the evolving nature of the virus. This study has the potential to provide valuable insights pertaining to the prevention of reinfection outbreaks and the associated health ramifications arising from reinfection occurrences.

## Data Availability

Individual, de-identified patient data can be available upon request from the corresponding authors.

## Abbreviations

COVID-19: Coronavirus disease 2019
SARS-CoV-2: Severe acute respiratory syndrome coronavirus 2
WHO: World Health Organization
PCR: polymerase chain reaction
RAT: rapid antigen test
SD: standard deviation
IQR: interquartile range
PSM: propensity score matching
aOR: adjusted odd ratios
OR: unadjusted odd ratios

## Declarations

### Ethics approval and consent to participate

Ethics committee of Hong Kong Baptist University waived ethical approval for this work. Informed consent for the telephone-visit data collection was provided by all patients. The reporting of this study is in accordance with the recommendations of the STROBE (Strengthening the Reporting of Observational Studies in Epidemiology) guidelines [37].

### Consent for publication

All authors meet the requirements for authorship as stated in this document and have read and approved the final manuscript.

### Availability of data and materials

Individual, deidentified patient data can be available upon request from the corresponding authors.

### Author contribution

L Tian, A Lyu, and Z Bian conceived and designed the study; Z Huang performed all the statistical analysis. Z Huang, J Luo, L Tian, A Lyu, and Z Bian interpreted the results. H Li and X Cheng contributed to the data processing; J Luo, J Zhang, H Wong, H To Tang, and C Cheung collected the raw data. Z Huang and J Luo wrote the manuscript; L Tian, A Lyu and Z Bian edited the manuscript. All authors read and approved the manuscript.

## Acknowledgements

We thank Leihan Tang, Linda Zhong, and Xin Xiong for valuable discussions.

