## Supplementary Information for "Incidence, risk factors, and clinical symptom profile of reinfection during Omicron-dominated COVID-19 outbreak in Hong Kong: A retrospective cohort study"

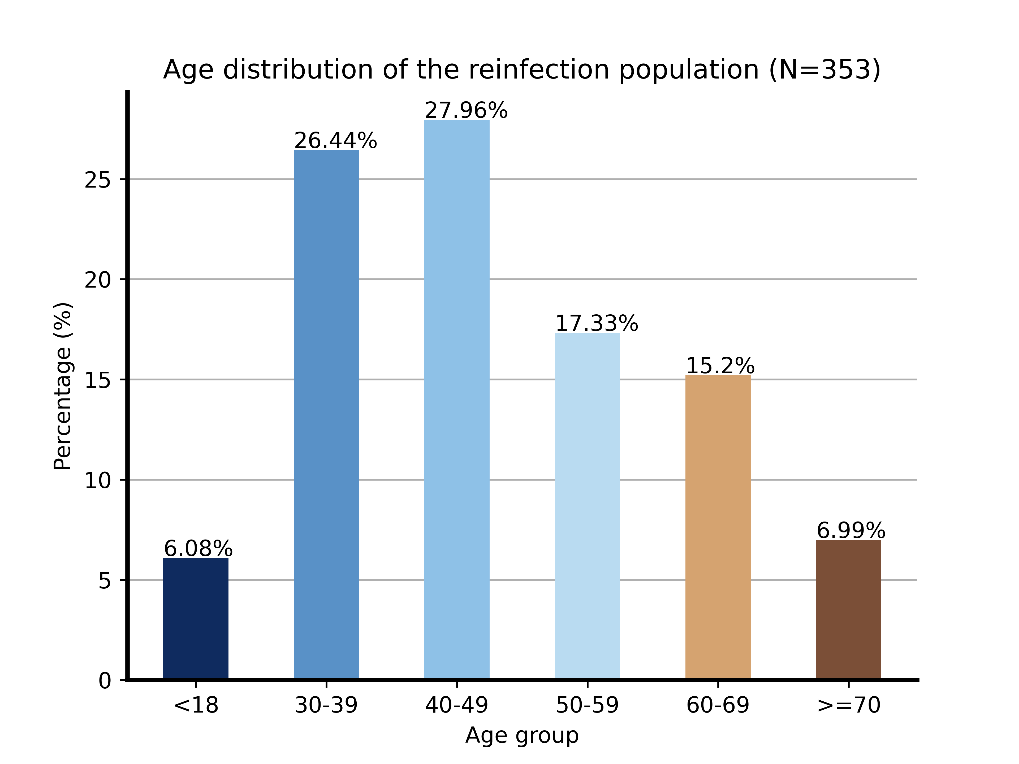

**Figure S1. Age distribution of the reinfection population (n = 353).**

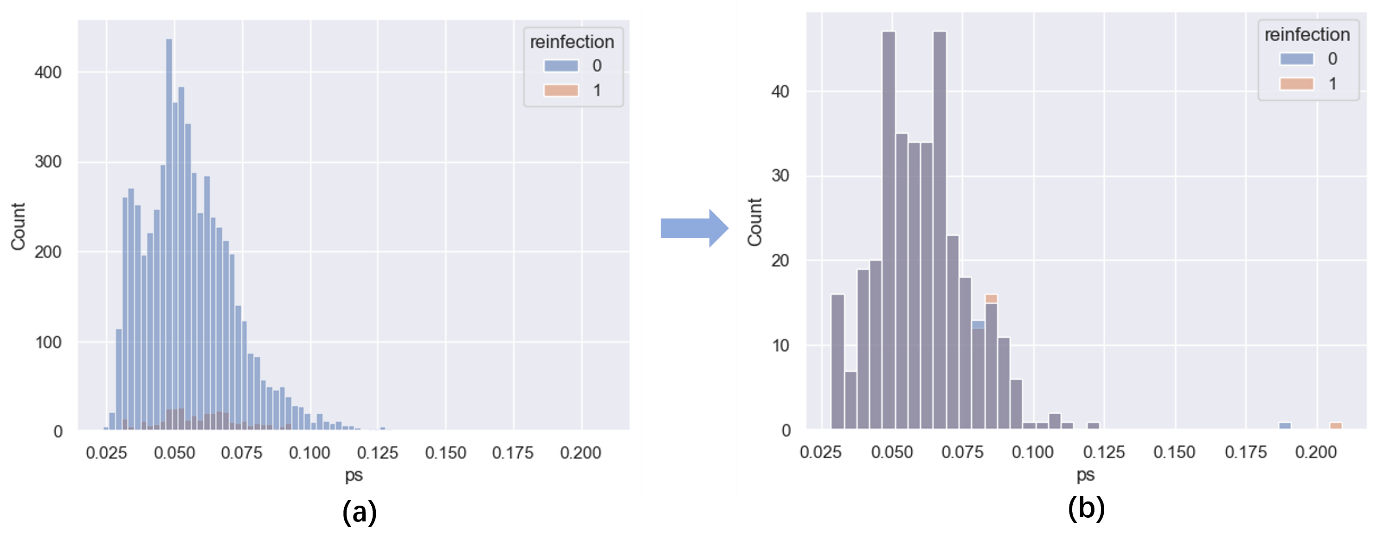

**Figure S2. Propensity score distributions of reinfection and non-reinfection groups.** (a) before matching: reinfection (n = 353) vs. non-reinfection (n = 5972); (b) after matching: reinfection (n = 353) vs. non-reinfection (n = 353).

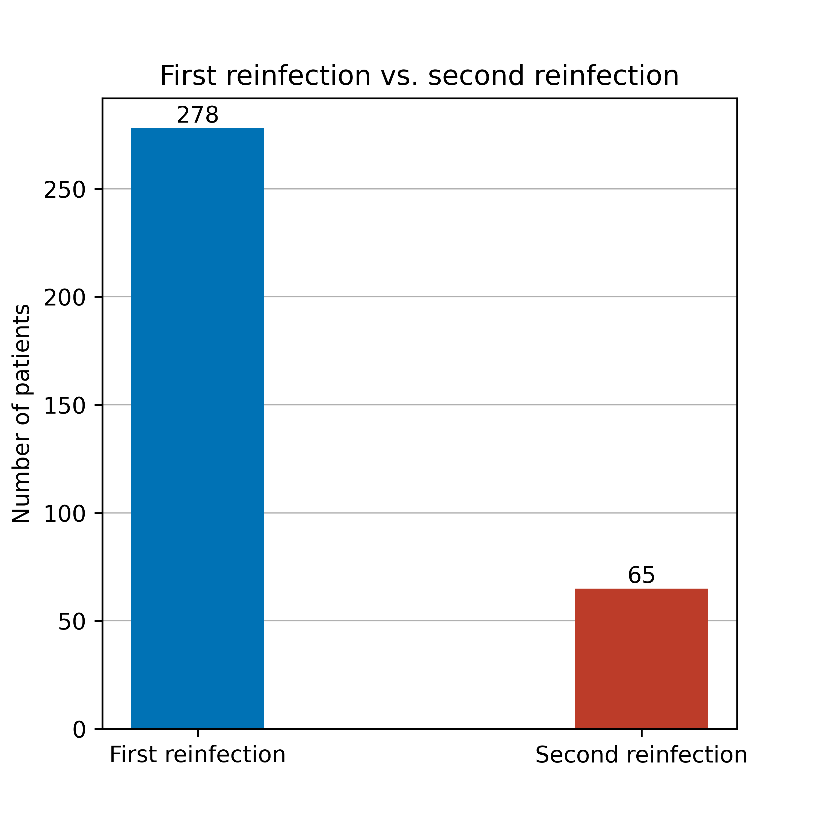

**Figure S3. Number of patients who have reinfected once vs. twice.** Among reinfection patients (n = 353), 78.75% (n = 278) of them had reinfection once, 21.25% (n = 65) had reinfection twice.

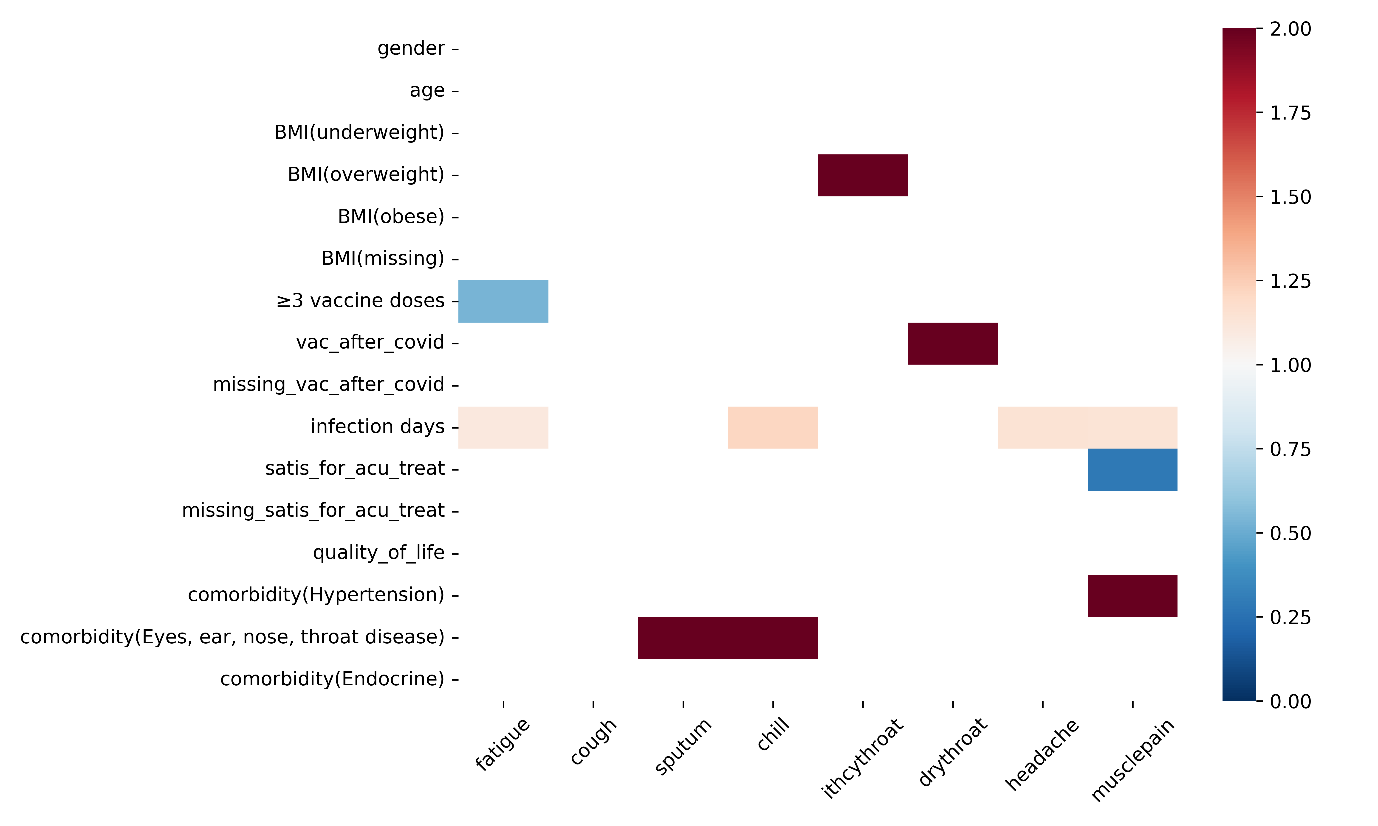

**Figure S4. Logistic regression matrix of the association between risk factors and top 8 most prevalent symptoms (>5%).** Individuals who experienced a single instance of reinfection were incorporated into the study, provided that the manifestation of symptoms occurred within a period not exceeding 90 days (≤ 90) from the occurrence of reinfection (N = 301). Each column contains adjusted odds ratio (aOR) of various factors computed by a multivariable logistic regression model for each symptom type. Only statistically significant aOR (p < 0.05) were presented, while non-significant aOR were blanked. *satis_for_acu_treat: satisfied with the treatments during the acute stage of primary infection; missing_satis_for_acu_treat: its missing values.

**Table S1. Types of new disease developed after primary COVID-19 infection**

| **New disease types** | 1. Hypertension |
| --- | --- |
|  | 2. Type-Ⅱ Diabetes |
|  | 3. Hyper |
|  | 4. Hyperlipidemia |
|  | 5. Cold |
|  | 6. Rhinitis |
|  | 7. Pneumonia |
|  | 8. Other respiratory diseases |
|  | 9. Endocrine disease (Thyroid Dysfunction) |
|  | 10. Liver disease |
|  | 11. Renal disease |
|  | 12. Digestive diseases (Gastritis, Enteritis, etc.) |
|  | 13. Cancer |
|  | 14. Neurological disease |
|  | 15. Other diseases |

**Table S2. Comparison of characteristics among first & second reinfection groups**

| **Characteristics** | **First reinfection (N = 278)** | **Second reinfection (N = 65)** | **P-value** |
| --- | --- | --- | --- |
| **Gender, n (%)** |  |  | 0.813 |
| Female | 216 (77.70%) | 62 (22.30%) |  |
| Male | 49 (75.38%) | 16 (24.62%) |  |
| **Age, median [IQR], years** | 41 (34, 52) | 44 (35, 58.75) | 0.109 |
| <18 years, n (%) | 15 (5.40%) | 4 (6.15%) | 1.000 |
| 18-29 years, n (%) | 20 (7.19%) | 3 (4.62%) | 0.636 |
| 30-39 years, n (%) | 63 (77.34%) | 23 (22.66%) | **0.049** |
| 40-49 years, n (%) | 76 (27.34%) | 15 (23.08%) | 0.586 |
| 50-59 years, n (%) | 42 (15.11%) | 12 (18.46%) | 0.632 |
| 60-69 years, n (%) | 43 (15.47%) | 6 (9.23%) | 0.273 |
| ≥ 70 years, n (%) | 19 (6.83%) | 2 (3.08%) | 0.395 |
| **BMI, median [IQR], kg/m2** | 23.9 (20.80, 26.00) | 22.7 (20.45, 25.85) | 0.166 |
| Underweight (<18.5), n (%) | 12 (4.32%) | 0 (0%) | 0.183 |
| Normal (18.5-23.0), n (%) | 75 (26.98%) | 13 (20.00%) | 0.316 |
| Overweight (23.0<25.0), n (%) | 26 (9.35%) | 7 (10.77%) | 0.908 |
| Obese (≥25.0), n (%) | 50 (17.99%) | 9 (13.85%) | 0.539 |
| **At least 3 doses of vaccine** | 124 (44.60%) | 31 (47.69%) | 0.755 |
| **Vaccination after infection** | 79 (30.27%) | 14 (24.56%) | 0.486 |
| **At least 1 comorbidity, n (%)** | 90 (32.37%) | 14 (21.54%) | 0.118 |
| Diabetes | 19 (6.83%) | 3 (4.62%) | 0.707 |
| Hypertension | 51 (18.35%) | 9 (13.85%) | 0.498 |
| Hyperlipidemia | 21 (7.55%) | 4 (6.15%) | 0.900 |
| Liver disease | 3 (1.08%) | 1 (1.54%) | 1.000 |
| Renal disease | 0 (0. %) | 1 (1.54%) | 0.428 |
| Psychological disease | 2 (0.72%) | 1 (1.54%) | 1.000 |
| Endocrine disease | 5 (1.80%) | 2 (3.08%) | 0.866 |
| Eyes, ear, nose, throat disease | 6 (2.16%) | 1 (1.54%) | 1.000 |
| Other disease | 12 (4.32%) | 3 (4.62%) | 1.000 |
| **Post-infection impact** |  |  |  |
| No impact | 103 (37.05%) | 37 (56.92%) | **0.005** |
| Acceptable impact | 113 (40.65%) | 18 (27.69) | 0.073 |
| Severe impact | 32 (11.51%) | 3(4.62%) | 0.154 |
| Missing | 30 (10.79%) | 7 (10.70%) |  |
